# An evaluation of community-based comprehensive vision care for the ageing population

**DOI:** 10.1101/2021.08.22.21262343

**Authors:** She Chiu Yang, Tsz Kin Law, Yan Lok Lucas Leung, Yim Ying Tam, Rita Sum, Jinxiao Lian, Maurice Yap

**Affiliations:** School of Optometry, The Hong Kong Polytechnic University

**Author notes:** **Corresponding author:** Dr Jinxiao Lian, School of Optometry, The Hong Kong Polytechnic University, Hung Hom, Kowloon, Hong Kong.

## Abstract

**Objective:** This study was conducted to evaluate the real-world effectiveness and potential cost-effectiveness of a community-based vision care programme for the ageing population aged 60 years or above.

**Methods:** Data from a total of 8899 subjects participating in a community based comprehensive vision care programme from 2015 to 2019 were extracted and analyzed to evaluate the effectiveness of the programme in terms of the prevalence of visual impairment (VI), the change in the prevalence of VI after refractive error correction, and the types of ocular disorders detected. VI was defined by a) visual acuity (VA) worse than 6/18 in any eye (worse eye) and b) VA worse than 6/18 in the better eye. The cost-effectiveness from the funder’s perspective was also estimated in terms of cost per VI avoided from such a programme.

**Results:** The study found that referenced to VA of the worse eye, the prevalence of VI was 39.1% (3482/8899, 95% CI: 38.1%-40.1%) based on presenting VA and reduced to 13.8% (1227/8899, 95% CI: 13.1%-14.5%) based on best-corrected VA. Referenced to the VA in the better eye, the prevalence of VI was 17.3% (1539/8899, 95% CI: 16.5%-18.1%) based on presenting VA and decreased to 4.2% (373/8899, 95% CI: 3.8%-4.6%) after best-corrected VA. Uncorrected refractive error was the major cause of presenting VI. From the funder’s perspective, the cost per VI case prevented was HK$1921 based on VA in the worse eye and HK$3715 based on the better eye.

**Conclusion:** A community-based programme was effective to detect the VI and reduced a relative 65% to 76% VI after refractive error correction for the community-dwelling ageing population. The ageing population can benefit from regular comprehensive vision care.

## Introduction

According to the World Health Organization (WHO), the proportion of the global population over 60 years old will increase from 12% to 22% between 2015 and 2050 ^1^. Ageing is associated with general functional loss and impairment, including vision ^2^. Age-related vision problems and disorders, such as uncorrected refractive error, cataract, age-related macular degeneration, and diabetic retinopathy, are thus expected to become more common in the population. Poor vision affects daily activities and quality of life and can result in negative consequences such as poor mental health, falls, and increased dependency ^3 4^. More people will be expected to have a visual impairment in a society with an ageing population. This points to the urgent need for healthcare and social systems to get ready for this demographic shift in the future.

Hong Kong (HK), from 2019 to 2050, the population of local elderly aged 60 years or above is estimated to increase from 25% to 40% ^5^. A population-based study among 3441 elders aged 60 years or above in Shatin district, a suburban area of HK, in 2002 found 41.3% of them with presenting visual acuity (VA) less than 6/18 in at least one eye ^6^. This percentage of visual impairment (VI) dropped to 34.5% when a pinhole was used. In addition, a proportion of 19.5% of subjects had presenting VA less than 6/18 in the better eye. The principal cause of VI was uncorrected refractive error, followed by cataract and macular degeneration. This study provided useful information to highlight the potential burden of VI among the elderly population 20 years ago. Since a pinhole was used to estimate the potential VA improvement, it could not simulate the effect of refractive correction with lenses. Over the ensuing 20 years, there were policy changes, improvements in a healthy lifestyle, and modernization of the health care service. All these may contribute to improved access to vision care and prevention of VI. A more recent cross-sectional study conducted between 2016 and 2018 found that the prevalence of VI (i.e. VA worse than 20/60 in the better eye) among HK elderly aged 60 years or above, who were recruited from the FAMILY cohort project, was 9.5% (63/661) and 2.0% (13/661) for presenting and best-corrected VA respectively ^7^. Similarly, uncorrected refractive error and cataract were the major causes of VI. However, this study targeted the adult population aged from 18 to 90 years, with low participation among older subjects, which may therefore underestimate the prevalence rate in the general population. The subjects in the study were recruited from a family cohort with some coming from the same family, which may have the potential of family clustering effect^7^.

WHO has called for global action to support healthy ageing by formulating evidence-based policies that strengthen the abilities of older persons and to align health systems with the needs of older populations ^1^. Older people who maintain good vision are more likely to maintain good functioning in activities of daily living, good social function, and independence. They are also more likely to avoid co-morbidities related to poor visual function such as falls and depression ^8^. A community-based programme, namely the “Kwai Tsing Signature Project Scheme” (KT programme), was established in 2015 in the Kwai Tsing district in HK, which provides comprehensive vision care to the community-dwelling ageing population. This provided an opportunity to evaluate the real-world evidence from a running community health service in terms of the effectiveness and potential cost-effectiveness of addressing the needs for vision care for an ageing population.

## Methods

### Community-based programme

This KT programme was launched by the Kwai Tsing District Council, in collaboration with the Kwai Tsing Safety Community and Healthy City Association, and the School of Optometry of The Hong Kong Polytechnic University (PolyU). This programme was publicized over a wide area by the District Council through various channels, such as advertisement and public event. Under the scheme, Kwai Tsing residents aged 50 or above could apply for a one-off comprehensive vision examination at the Integrative Community Health Centre (ICHC) in Lai King with a copayment of HK$15 (HK$10 initially during 2015-2017) and purchase a pair of single-vision spectacles with a copayment of HK$75 (HK$50 initially during 2015-2017) if indicated. The District Council, with funding from the Home Affairs Bureau of the Hong Kong SAR government, subsidized HK$250 for each examination and HK$550 (HK$500 initially during 2015-2017) for a pair of prescription spectacles. Those residents who were under the Comprehensive Social Security Assistance (CSSA) Scheme or recipients of fringe benefits for civil servants and their relatives were not eligible for this KT programme. The comprehensive vision examination was conducted by optometrists in the ICHC of PolyU following the routine optometric examination protocol, including history taking, VA measurement and preliminary testing, objective and subjective refraction, external and internal ocular health assessment and additional tests if indicated.

### Subjects and data extraction

All subjects who have participated in the KT programme, aged 60 years or above, and with complete examination records were eligible for this study. Their examination records were extracted, including demographic data, entrance VA, best-corrected VA (BCVA), and the types of suspected ocular disorders. VA was classified into four levels by two definitions, i.e., VA based on the a) worse eye or b) better eye. Based on the worse eye, “no VI” was defined as VA of 6/18 or better in both eyes, “moderate VI” as VA worse than 6/18 to 6/60 in the worse eye, “severe VI” as VA worse than 6/60 to 6/120 in the worse eye, and “blindness” as VA worse than 6/120 (Table 1)^9^. Prevalence of any VI was defined as having at least one eye with VA less than 6/18. Based on the better eye b) the levels of VI was defined based on the VA in the better eye following the same criteria as above to define no, moderate, severe VI and blindness. Prevalence of any VI was defined as having better eye VA less than 6/18, as the definition study by Michon et al ^6^. The suspected ocular disorders were classified into 3 categories: uncorrected refractive error, anterior disorders such as cataract and corneal scars, and posterior disorders such as macular hole, age-related macular degeneration and epiretinal membrane.

**Table 1.**
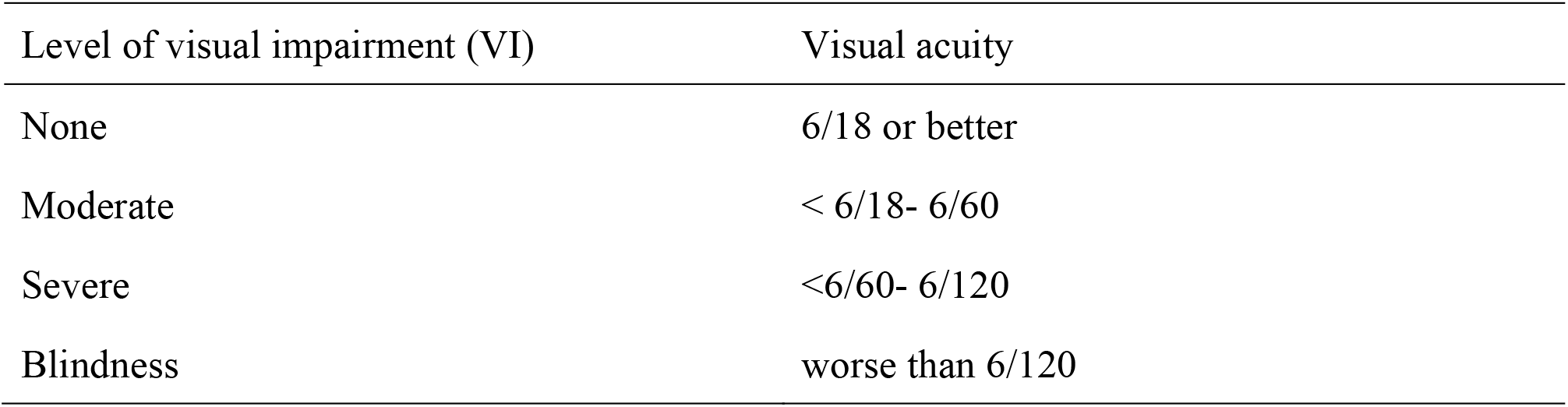
Definition of visual impairment.

### Statistical analysis

Descriptive analyses were used to summarize the prevalence of VI with a 95% confidence interval (CI), the changes in the levels of VI after refractive error correction, and the suspected ocular disorders. The prevalence of VI was compared between sex, and age groups using the Chi-square test. SPSS statistical software version 26.0 was used for the analyses.

## Results

From 2015 to 2019, a total of 9149 subjects participated in the KT programme, but 250 subjects not aged 60 years or above or with missing or invalid data were excluded, leaving 8899 subjects eligible for the analysis. Among them, 61.9% (5507/8899) were female participants and 46.7% (4159/8899) were 70 years or above (Table 2).

**Table 2.** Demographic characteristics of the subject group by sex and age (N=8899)

| **Parameters** | **n (%)** |
| --- | --- |
| **Sex** |  |
| Male | 3392 (38.1) |
| Female | 5507 (61.9) |
| **Age** |  |
| 60-64 | 1813 (20.4) |
| 65-69 | 2927 (32.9) |
| 70-74 | 2055 (23.1) |
| 75 or above | 2104 (23.6) |

### Prevalence of visual impairment

The overall prevalence of VI (using the definition of worse than 6/18 in at least one eye) in the elderly aged 60 or above was 39.1% (3482/8899, 95% CI: 38.1%-40.1%) based on presenting VA and 13.8% (1227/8899, 95% CI: 13.1%-14.5%) based on BCVA respectively (Table 3). The largest reduction was the prevalence of moderate VI which reduced from 33.0% (2937/8899) based on presenting VA to 10.4% (925/8899) based on BVCA, followed by the prevalence of severe VI which reduced from 3.7% (329/8899) to 1.4% (125/8899), and lastly in the group with blindness from 2.4% (216/8899) to 2.0% (177/8899).

**Table 3.** Prevalence of visual impairment based on presenting VA and BCVA.

| Level of visual impairment (VI) | Prevalence of visual impairment, n (%), N=8899 |  |  |  |
| --- | --- | --- | --- | --- |
|  | VA less than 6/18 in at least one eye |  | VA less than 6/18 in the better eye |  |
|  | Based on presenting VA | Based on BCVA | Based on presenting VA | Based on BCVA |
| None ($\leq 6/18$) | 5417 (60.9) | 7672 (86.2) | 7360 (82.7) | 8526 (95.8) |
| Moderate ($> 6/18$ and $\leq 6/60$) | 2937 (33.0) | 925 (10.4) | 1480 (16.6) | 359 (4.0) |
| Severe ($> 6/60$ and $\leq 6/120$) | 329 (3.7) | 125 (1.4) | 49 (0.6) | 11 (0.1) |
| Blindness ($> 6/120$) | 216 (2.4) | 177 (2.0) | 10 (0.1) | 3 (0.03) |
| Sub-total of visually impaired | 3482 (39.1) | 1227 (13.8) | 1539 (17.3) | 373 (4.2) |

The overall prevalence of VI (using the definition of worse than 6/18 based on the better eye) was 17.3% (1539/8899, 95% CI: 16.5%-18.1%) based on presenting VA and reduced to 4.2% (373/8899, 95% CI: 3.8%-4.6%) based on BCVA.

There was more VI in older age groups. Comparing the 60-64 group to the group aged 75 or above, VI increased from 22.5% (407/1813) to 62.1% (1306/2104), based on VA of the worst eye, and from 8.4% (152/1813) to 32.9% (693/2104), based on VA of the better eye (P-value<0.001, Table 4).

**Table 4.** Prevalence of visual impairment based on presenting VA by sex and age group.

| Parameters | Have visual impairment, n (%) | No visual impairment, n (%) | Total | P-value* |
| --- | --- | --- | --- | --- |
| **Visual impairment based on the worse eye (N=8899)** |  |  |  |  |
| Sex |  |  |  | 0.041 |
| Male | 1373 (40.5) | 2019 (59.5) | 3392 (100.0) |  |
| Female | 2109 (38.3) | 3398 (61.7) | 5507 (100.0) |  |
| Age |  |  |  | <0.001 |
| 60-64 | 407 (22.5) | 1406 (77.6) | 1813 (100.0) |  |
| 65-69 | 920 (31.4) | 2007 (68.6) | 2927 (100.0) |  |
| 70-74 | 849 (41.3) | 1206 (58.7) | 2055 (100.0) |  |
| 75 or above | 1306 (62.1) | 798 (37.9) | 2104 (100.0) |  |
| **Visual impairment based on the better eye (N=8899)** |  |  |  |  |
| Sex |  |  |  | 0.588 |
| Male | 596 (17.6) | 2796 (82.4) | 3392 (100.0) |  |
| Female | 943 (17.1) | 4564 (82.9) | 5507 (100.0) |  |
| Age |  |  |  | <0.001 |
| 60-64 | 152 (8.4) | 1661 (91.6) | 1813 (100.0) |  |
| 65-69 | 334 (11.4) | 2593 (88.6) | 2927 (100.0) |  |
| 70-74 | 360 (17.5) | 1695 (82.5) | 2055 (100.0) |  |
| 75 or above | 693 (32.9) | 1411 (67.1) | 2104 (100.0) |  |
| * Chi-square test |  |  |  |  |

Among subjects with VI based on the worse eye, the proportion of correctable vision from any VI to no VI was significantly higher in the younger age group than the older groups, reduced from 79.6% (324/407) in the 60-64 aged group to 50.0% (653/1306) in the 75 or above aged group (P-value<0.001, Table 5). When the VI was defined based on the better eye, 92.1% (140/152) VI can be correctable in the aged group 60-64 and 64.1% (444/693) in the aged group 75 or above.

**Table 5.** Proportion of correctable visual impairment by sex and age group.

| Parameters | Correctable visual impairment, n (%) | Uncorrectable visual impairment, n (%) | Total | P-value* |
| --- | --- | --- | --- | --- |
| **Visual impairment based on the worse eye (n = 3482)** |  |  |  |  |
| Sex |  |  |  | 0.020 |
| Male | 857 (62.4) | 516 (37.6) | 1373 (100.0) |  |
| Female | 1398 (66.3) | 711 (33.7) | 2109 (100.0) |  |
| Age |  |  |  | <0.001 |
| 60-64 | 324 (79.6) | 83 (20.4) | 407 (100.0) |  |
| 65-69 | 702 (76.3) | 218 (23.7) | 920 (100.0) |  |
| 70-74 | 576 (67.8) | 273 (32.2) | 849 (100.0) |  |
| 75 or above | 653 (50.0) | 653 (50.0) | 1306 (100.0) |  |
| **Visual impairment based on the better eye (n = 1539)** |  |  |  |  |
| Sex |  |  |  | 0.755 |
| Male | 449 (75.3) | 147 (24.7) | 596 (100.0) |  |
| Female | 717 (76.0) | 226 (24.0) | 943 (100.0) |  |
| Age |  |  |  | <0.001 |
| 60-64 | 140 (92.1) | 12 (7.9) | 152 (100.0) |  |
| 65-69 | 289 (86.5) | 45 (13.5) | 334 (100.0) |  |
| 70-74 | 293 (81.4) | 67 (18.6) | 360 (100.0) |  |
| 75 or above | 444 (64.1) | 249 (35.9) | 693 (100.0) |  |
| * Chi-square test |  |  |  |  |

### Suspected ocular disorders among uncorrectable visual impairment

Ocular disorders were classified into anterior and posterior types. Among 3482 subjects with VI with presenting VA defined based on worse eye, 2255 had correctable refractive error, the remaining were still visually impaired in at least one eye after refractive error correction, including 612 subjects with anterior disorders only, 235 with posterior disorders only, 146 with both anterior and posterior disorders and 234 subjects with unknown causes (Table 6).

**Table 6.**
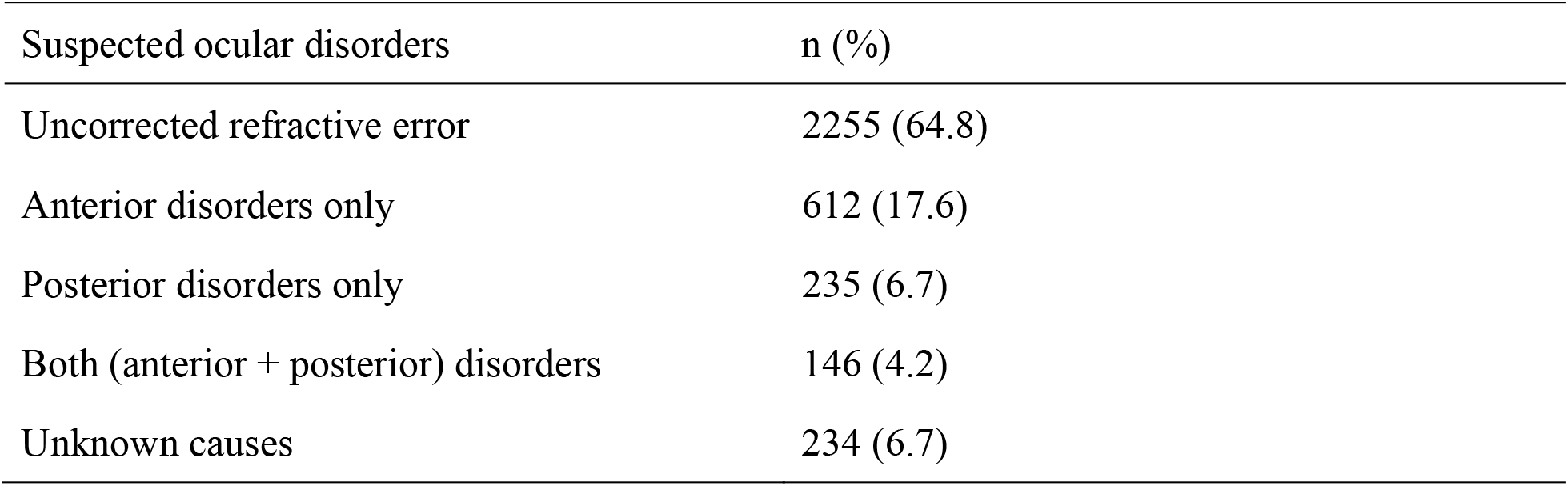
Suspected ocular disorders for those who have impairment in any eye (n = 3482)

### Cost-effectiveness

The latest subsidized amount for each subject receiving the examination was HK$250, and a co-payment of HK$15 was paid by each subject. The subsidized amount for each prescribed pair of glasses was HK$550, with a co-payment of HK$75 from each subject receiving the glasses. Using the data of 8899 subjects from this community service programme, from the funder’s perspective, the total cost of subsidizing the examination was HK$2,091,265 ((HK$250-HK$15)*8899) and the total cost of subsidizing the glasses is HK$2,240,323 ((HK$550-HK$75)*8899*53%) by a proportion of around 53% required the new prescription of glasses for refractive error correction, which results in the total of HK$4,331,588. After the refractive error correction (Table 3), a total number of 2255 subjects improved from any VI to no VI using a) VA based on the worse eye and a total of 1166 subjects using b) VA based on the better eye. Therefore, the cost per VI avoided from this community-based programme was estimated to be HK$1921 based on the worse eye and HK$3715 based on the better eye from the funder’s perspective.

## Discussion

This study evaluated the effectiveness and potential cost-effectiveness of a community-based programme to detect and address the vision problem for the community-dwelling ageing population. The results provide real-world evidence which would be useful for service planning for vision care for the ageing population.

This government-funded programme was initiated by a district council and was widely publicized in the relevant district. The participating elders represent those who would normally respond to public health interventions in their community. This programme was provided at a very low co-payment cost to the elders (HK$15=US$1.9) for the examination and HK$75 (US$=9.6) for a pair of glasses, so there was a minimal financial barrier. Although the participating subjects might not be representative of the whole elderly population in HK, we expect that they represented a more health-conscious group. From this community-based programme, a high prevalence of VI was found. Nearly one in three elders had at least one eye with VI (VA based on the worse eye: 39.1%) and about 1 in 6 elders had both eyes with VI (VA based on the better eye: 17.3%). With refractive error correction, 65% of those with VI (from 39.1% to 13.8%) with presenting VA based on the worse eye was improved to no VI and it was 76% of VI improved to no VI with VA based on the better eye (from 17.3% to 4.2%). This significant drop in the proportion of VI suggests that a majority of the elders with VI can benefit from this type of community-based programme from which VI can be detected and addressed with refractive error correction immediately, especially among the groups with moderate to severe VI and the younger aged groups.

The prevalence of VI from this community-based programme was similar to one population-based study conducted in another district (Shatin) in HK 20 years ago which reported a 41.3% prevalence of VI with VA worse than 6/18 in at least one eye^6^. It is worth noting that the sampling methods were different; the Shatin study was a population-based study using cluster sampling while a convenient sampling based on participants in a government-funded community-based programme was used in our study. Although using different sampling methods, both studies ended up with a similar demographic profile of the elders (mean age of 70.2 years, females 61.9%, and males 38.1%) in our study and those in the Shatin study (mean age of 70.2 years in men and 70.6 years in women; females 60.0% and males 40.0%). It is worth noting that pinhole measurement instead of subjective refraction was used in the Shatin study. When pinhole VA was used, the prevalence of VI was reduced from 41.3% with presenting VA to 34.5% with pinhole, only with a 16.5% relative reduction in the VI from the Shatin study. The pinhole reduces the influence of spherical aberration and diffractive effects of media opacities. An improvement in VA with a pinhole does not imply a refractive error ^10^. The pinhole acuity may also be several letters worse than the subjective BCVA^11^.

A recent 2020 cross-sectional study in HK reported the prevalence of VI with presenting VA worse than 6/18 in the better eye to be 9.5% among those aged 60 years or above^7^. Using the same definition of VI, our study reported a higher prevalence of VI (17.3%). As mentioned by the authors in the latest study, a low proportion of older elders (i.e., 39.2% aged 70 years or above among those aged 60 years or above) were recruited which may therefore underestimate the prevalence rate. Our study has a proportion of 46.7% (4159/8899) elders aged 70 years or above. A similar magnitude of 78.9% reduction in the VI after refractive error correction (from 9.5% to 2.0%) was reported in that latest study.

Among the 1227 subjects who were still visually impaired after refractive correction in this community-based programme, the most common suspected ocular disorder for VI was anterior disorders including cataract, pterygium, corneal degenerations and dystrophies, and posterior disorders including age-related macular degeneration, myopic maculopathy, epiretinal membrane, diabetic retinopathy, glaucoma and macular hole. However, these were the cases that could potentially benefit from the referral to specialists care for close monitoring and timely management.

One limitation of this study was that no data was collected on individual socioeconomic status, knowledge about ageing and vision problems, or other health records such as chronic diseases. Therefore, we were not able to perform further data analysis to determine the factors associated with the VI. Moreover, the scheme excluded applicants receiving CSSA and fringe benefits (such as civil servants). This could also lead to a selection bias and hence underestimation of the prevalence of VI if the excluded group were associated with a higher risk of VI, especially the group receiving CSSA. Besides these limitations, other limitations had been mentioned in previously published studies. For example, when defining VI, central visual acuity was the only focus, which did not consider the effect of visual field size. This might be a problem encountered by glaucoma patients, in which they experience progressive visual field loss and constriction at an advanced stage even though VA may still be fair ^12^. However, it is possible that some participants could have good central vision but poor peripheral visual fields in reality. As a result, current studies may not reflect the true prevalence in the elderly population. Nevertheless, as most previous studies also focused on central vision only, it was still possible for us to compare our results with other studies.

This is a large-scale community-based programme providing vision care to over 8000 elders. We estimated that it cost from HK$1921 to HK$3715, from the funder’s perspective, to prevent one VI case. Elders can benefit from such a community-based programme which would potentially result in better daily physical function, quality of life and independence by avoiding VI. The findings from this study provide a quantifiable basis to consider whether or not to provide subsidized vision care for the elderly. Future research should look holistically at the societal level, the cost-benefit of minimizing VI in the ageing population.

## Conclusion

The prevalence of VI was relatively high among older people in HK. The main cause was uncorrected refractive errors. Having access to regular vision care and affordable prescription spectacles would reduce the prevalence of VI and associated risks.

## Data Availability

Data not available due to ethical restrictions.

## Acknowledgements

We thank Mr Chow Yick Hay, Kwai Tsing Safety Community and Healthy City Association, for initiating this vision care programme and for supporting our study. Maurice Yap is supported by the KB Woo Family Endowed Professorship in Optometry.

